# Prevalence of Long COVID symptoms in Bangladesh: A Prospective Inception Cohort Study of COVID-19 survivors

**DOI:** 10.1101/2021.07.03.21259626

**Authors:** Mohammad Anwar Hossain, K M Amran Hossain, Karen Saunders, Zakir Uddin, Lori Maria Walton, Veena Raigangar, Mohamed Sakel, Rubayet Shafin, Mohammad Sohrab Hossain, Md. Feroz Kabir, Rafey Faruqui, Shohag Rana, Md. Shahoriar Ahmed, Sonjit Kumar Chakrovorty, Md. Anwar Hossain, Iqbal Kabir Jahid

## Abstract

**Objective:** The objective of this study was to identify the prevalence of ‘Long COVID’ symptoms (LCS) in a large cohort of survivors and identify any potential associated risk factors.

**Design:** A prospective survey was undertaken of an inception cohort of confirmed COVID-19 survivors (Aged 18 to 87 years).

**Participants and Setting:** 14392 participants were recruited from 24 testing facilities across Bangladesh between June, and November 2020. All participants had a previously confirmed positive COVID-19 diagnosis, and reported persistent symptoms and difficulties in performing daily activities.

**Main Outcome Measures:** Participants who consented, were contacted by face-to-face interview, and were interviewed regarding LCS, and restriction of activities of daily living using Post COVID-19 functional scale. Cardio-respiratory parameters were also measured.

**Results:** Among 2198 participants, the prevalence of LCS at 12 weeks was 16.1%. Overall, eight LCS were identified and in descending order of prominence are: fatigue, pain, dyspnea, cough, anosmia, appetite loss, headache, and chest pain. COVID survivors experienced between 1 to 5 LCS with an overall duration period of 21.8 ± 5.2 weeks. SEM predicted the length of LCS to be related to younger age, female gender, rural residence, prior functional limitation and smoking.

**Conclusion:** In this cohort of survivors, at 31 weeks post diagnosis, the prevalence of LCS was 16.1%. The risk factors identified for presence and longer length of LCS warrant further research and consideration to support public health initiatives.

## Introduction

Individual recovery from COVID-19 infection varies, and it is not fully understood why some people experience persistent symptoms over a longer time period than others. Post-acute COVID-19 syndrome^1^ (PACS)and Post-COVID-19 syndrome^2^ are terms used clinically to describe ongoing or new symptoms that occur after the fourth week of recovery from an acute infection, that cannot be explained by an alternative medical diagnosis. The experience of longer-term symptoms has prompted some patient groups to invent and use the term “Long COVID”^3^ to describe their experience of ongoing symptoms, that persist beyond four weeks from suspected infection or positive diagnosis to over 12 weeks and longer. It is now understood that COVID-19 can impact on multiple organ systems^4^, which can lead to a diverse range of persistent symptoms including fatigue, breathlessness, cough, loss of taste and/or smell, myalgia, memory issues and gastrointestinal problems.^5-6^An integrative post COVID symptom model was recently proposed for symptom classification after confirmed diagnosis of COVID-19^7^This model incorporates reference to the “relapsing remitting”^8^pattern of symptoms experienced by many survivors and has been adopted to assist in presentation of results.

A large recent survey conducted over a four-week period in the UK found that 1.1 million people self-reported “long COVID” symptoms (LCS), which equates to 1.7% of the population.^9^ The survey also identified that 18.1% of participants reported that their daily activities had been affected a lot by the illness, indicating that “Long COVID” has a detrimental impact on peoples’ day to day lives, so should be taken seriously.^10^

It is acknowledged that there is a need for further research into the prevalence and duration of LCS experienced by survivors^11^ and potential associated risk factors so that overall clinical management can be improved^12^.This is especially pertinent in Low to Middle Income Countries^13^ like Bangladesh, where the majority of the population live in rural districts^14^outside of the densely populated capital city, Dhaka. There are only two published studies so far in Bangladesh on persistent symptoms following COVID-19 infection. One survey of 1002 individuals reported that 20% had experienced persisting symptoms after COVID-19, with diarrhea (12.7%) being most common followed by fatigue (11.5%).^15^ A second smaller study of 355 individuals found that 46% of patients recovering from COVID-19 reported LCS^16^with fatigue being the most common symptom reported. Therefore, it is critical to gain research knowledge on the prevalence of LCS and identify associated risk factors, as this study does, which will be of relevance to the global community.

The aim of this study was to identify the prevalence of LCS in Covid-19 survivors and explore any potential associations between reported symptoms and the independent variables measured. Cardio-respiratory parameters and functional restrictions were also measured to try to discern if there was an impact on the cardiac system and if functional daily activities were affected by LCS.

## Methods

### Study design and participants

This prospective study utilized an inception cohort of adult COVID-19 survivors recruited from a large population sample frame of 14392 COVID positive cases. These cases were identified from 24 testing facilities across Bangladesh between June and November 2020. All COVID-19 positive and negative diagnoses were performed using a RT-PCR test (Real time polymerase chain reaction test)^17^. Inclusion criteria were: age 18 years and over; people who reported persistent symptoms after positive diagnosis and people who reported difficulties in undertaking usual daily activities. Exclusion criteria were: individuals too sick to participate; those who declined consent and those we were unable to contact.

The sample size calculation was performed using “EPI INFO” software version 7.4.2.0 developed by the Center for Disease Control in the US. For the calculation, the reference figure of 535,139 was used (ie. The total number of COVID-19 positive cases reported up to January 2021)^18^ with a cluster figure of eight (the number of administrative divisions in Bangladesh) A calculation was then made with 50% of expected frequency, 5% margin of error, and 1.0 design effect. The sample size was generated as a minimum of 1088 with a minimum of 136 samples per division.

### Study procedure

A clear flow diagram of the study process has been produced in Figure 1 to meet the quality guidelines recommended by Strengthening the Reporting of Observational studies in Epidemiology (STROBE)^20^.

**Figure 1:**
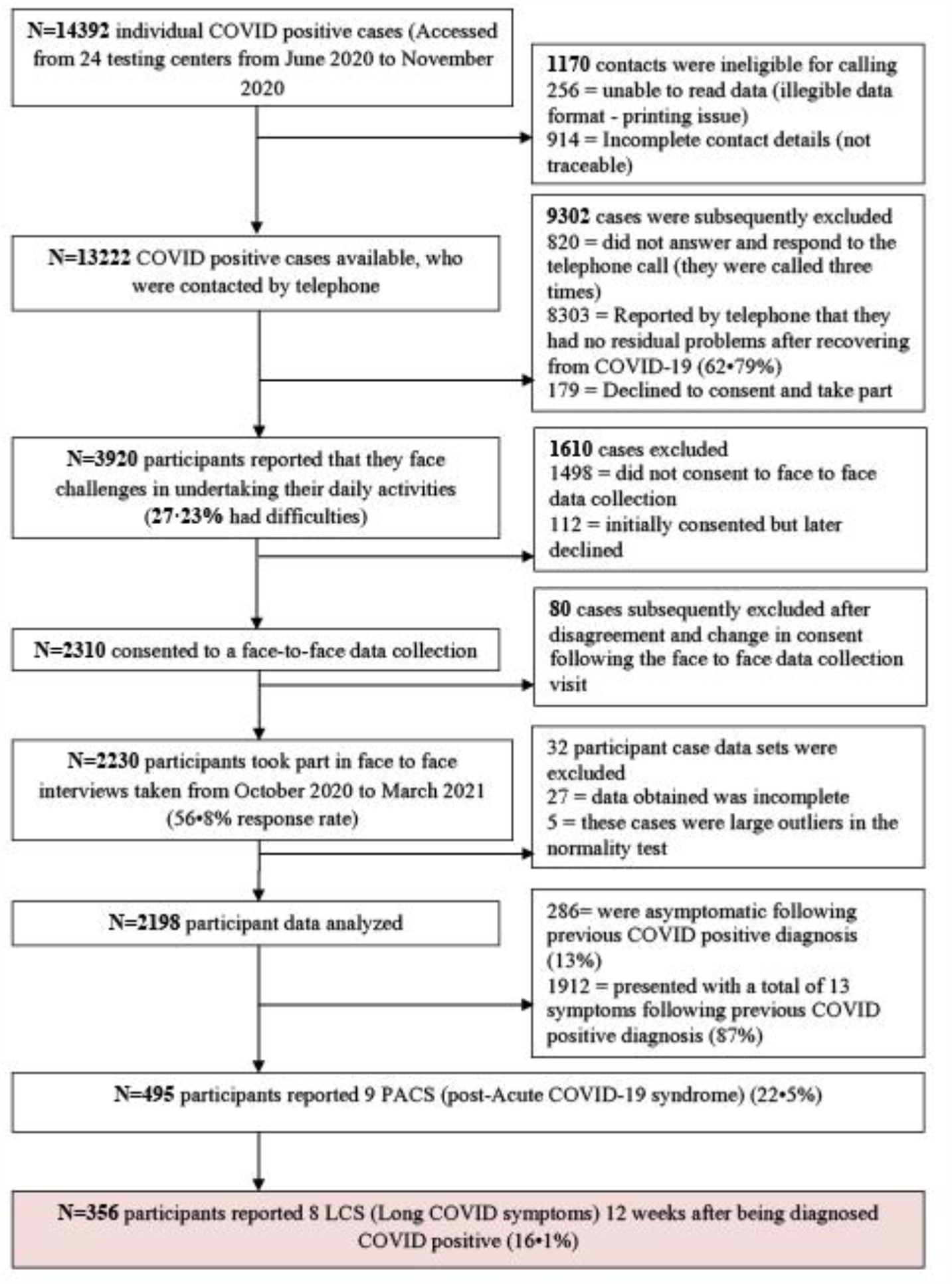
STROBE flow diagram of the study

**Figure 2:**
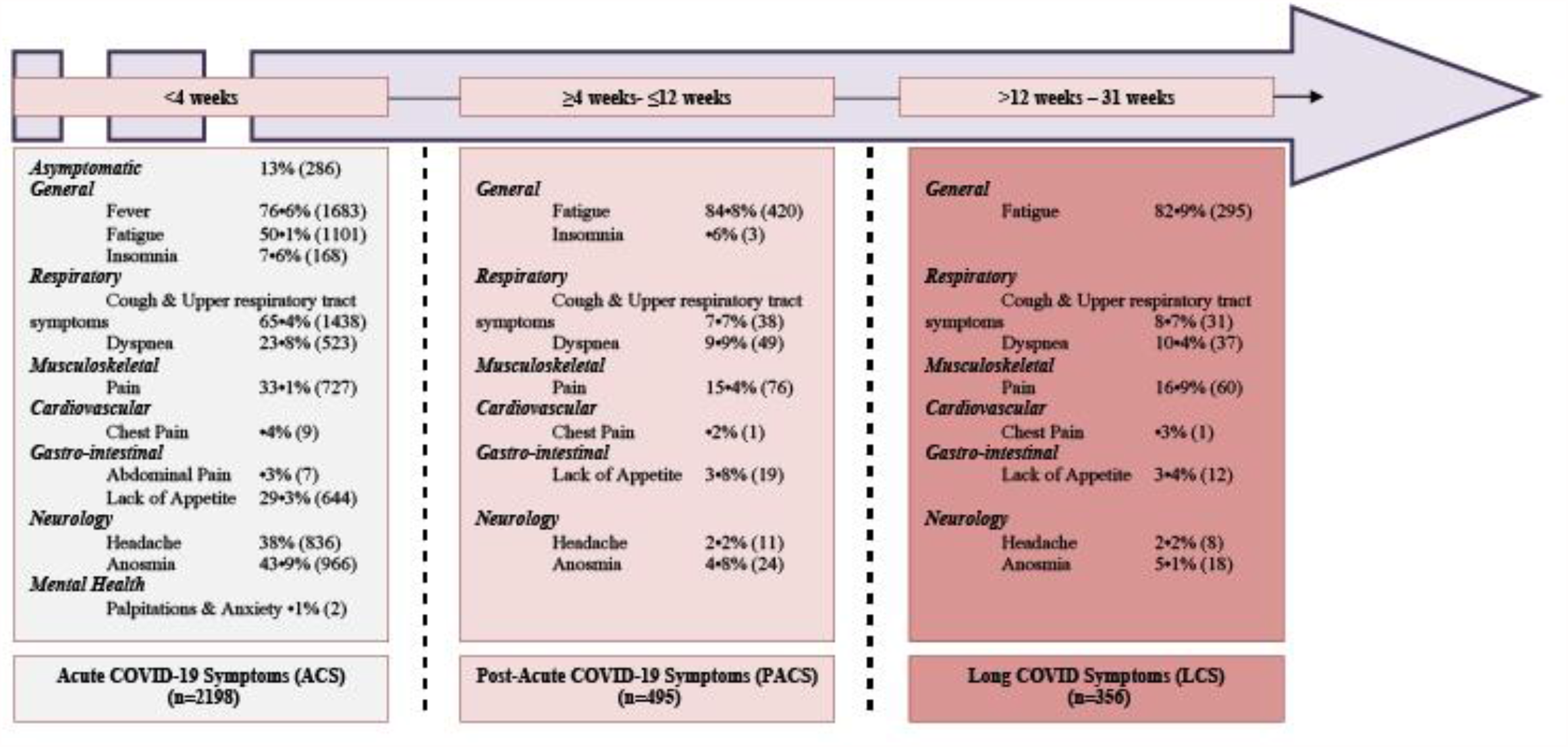
Acute COVID-19 Symptoms (ACS), Post-Acute COVID-19 symptoms (PACS) and Long COVID Symptom (LCS) responses

Data was collected by eight trained data collectors from the Centre for the Rehabilitation of the Paralyzed (CRP). Data collectors were comprehensively trained by the study team regarding study aims, ethical considerations, questionnaires and clinical outcome measures. Screening for trial eligibility and consent for data collection was conducted using mobile telephones. Face to face data collection was undertaken via mutually convenient scheduled appointments either in the respondent’s house or workplace. All data collectors adhered scrupulously to the COVID-19 preventative precautions, use of personal protective equipment (PPE) and health regulations. Consent and questionnaire documents were provided as hard paper copies in Bengali for respondents to complete themselves. If there was a literacy issue then data collectors provided support with this to enable completion. Data was then collated anonymously from these documents and transferred into an Excel workbook for audit and analysis. A small-scale pilot study using 17 respondents was initially conducted to test applicability and feasibility of the questionnaire.

### Data collection and Questionnaire

All contactable participants (N = 13,222) were asked by telephone: “Do you feel that you have any challenges or have any persisting symptoms after receiving a negative test result for COVID-19?” Participants who consented to face to face data collections (N = 2310) were provided with a questionnaire consisting of three parts. The first part was designed to gather socio demographic information with seven questions related to age, gender, marital status, education, residing area, and occupation. The second part consisted of seven questions related to comorbidities, blood group and rhesus status, date of COVID-19 positive test, date of COVID-19 negative test, presenting symptoms during COVID-19 illness, persisting COVID-19 symptoms, and treatment received during COVID-19 illness. The third part of the questionnaire focused on the measurement of cardio-respiratory parameters and included the Post COVID-19 Functional Status Scale^21^ (PCFS). Cardio-respiratory parameters measured included: resting heart rate (HR); blood oxygen saturation levels (Spo2); systolic and diastolic blood pressure; inspiratory and expiratory lung volumes; and maximal oxygen consumption (Vo2max).

The PCFS is an instrument that aims to identify and record the course of symptoms following infection with COVID-19 and their impact on the abilities of the recovering individual. The scale covers six domains including: survival; constant care; basic activities of daily living; instrumental activities of daily living; participation in usual social roles and a symptom checklist. Each item in each domain is scored from five possible options on an ordinal scale from zero to four with the fifth grade being “Death”. An overall final scale grade is obtained from completion with a high-grade corresponding to more functional limitations and a low final grade indicates no persisting symptoms or restricted daily activities. The PCFS has adequate construct validity^22^and has a Cronbach alpha score of α 0.879 in our study, which indicates a satisfactory level of internal consistency. The PCFS was translated into Bengali and the language validation process was followed as per WHO guidance.^23^

### Statistical Testing

Data analysis utilized the statistical software package for social sciences (SPSS) version 20.0. The normality test was performed through Kolmogorov-Smirnov test. Descriptive statistics were performed separately for Acute COVID-19 Symptoms (ACS), PACS and the LCS groups (Tables 1 and 2). Relationships between a categorical independent variable (for example, LCS) and a parametric socio-demographic dependent variable were determined through independent t tests. In addition, relationships among two or more categorical variables were explored using the chi-square test (Table 2). To ascertain which factors were potentially related to LCS, binary logistic regression was performed with the presence of LCS as the dependent variable (Table 3) and multiple linear regressions were performed with the duration of LCS as the dependent variable (Table 4). Figures 3, 5 and 6 are presented as bar charts with an error bar (95% CI). Figure 7 presents the associated risk factors identified for LCS using Structural Equation Modeling (SEM) using SPSS AMOS version 24.0 (Figure 7). The alpha value was set as p<.05.

**Table 1:**
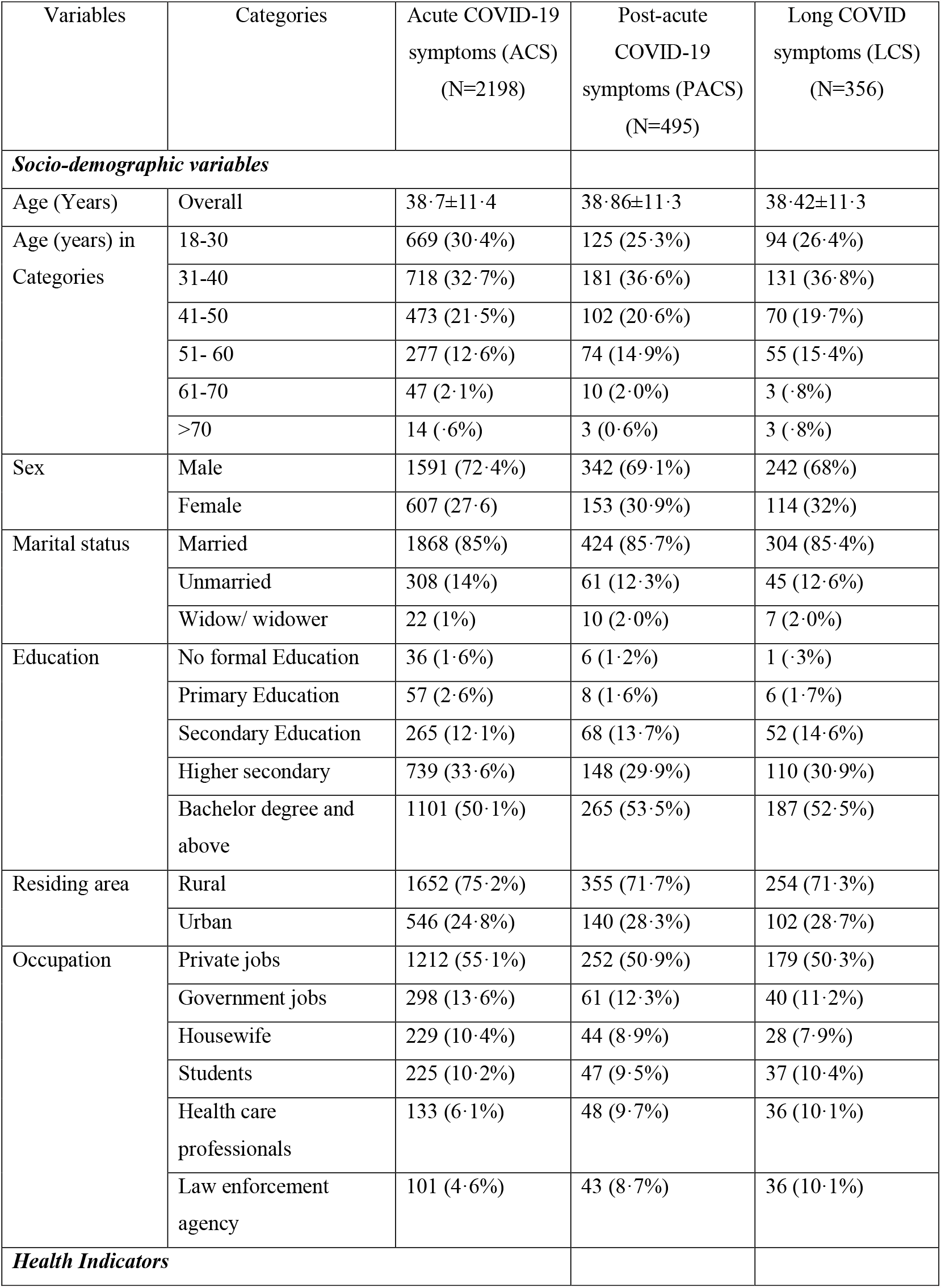
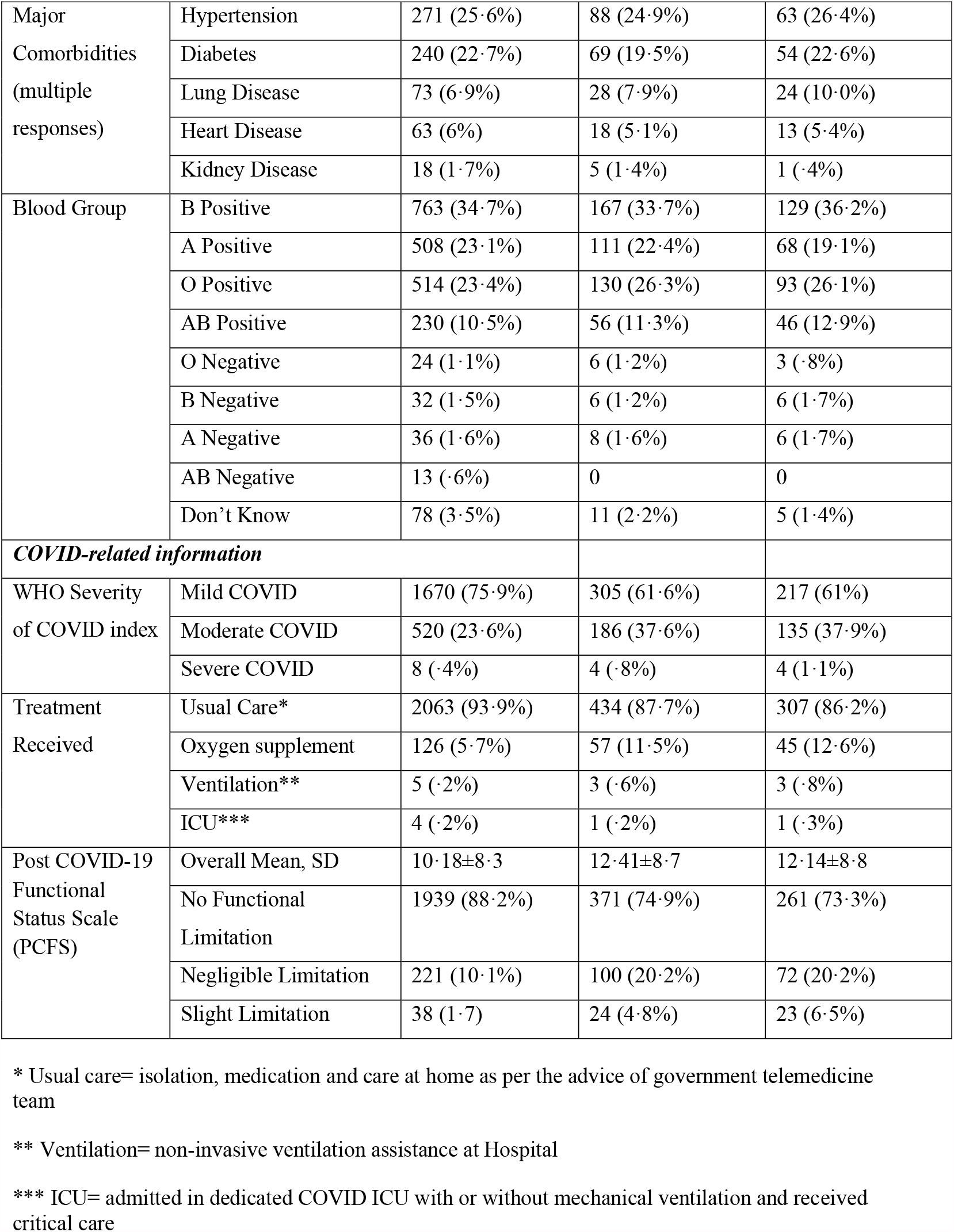
Baseline characteristics for Inception Cohort of COVID-19 positive cases

**Table 2:**
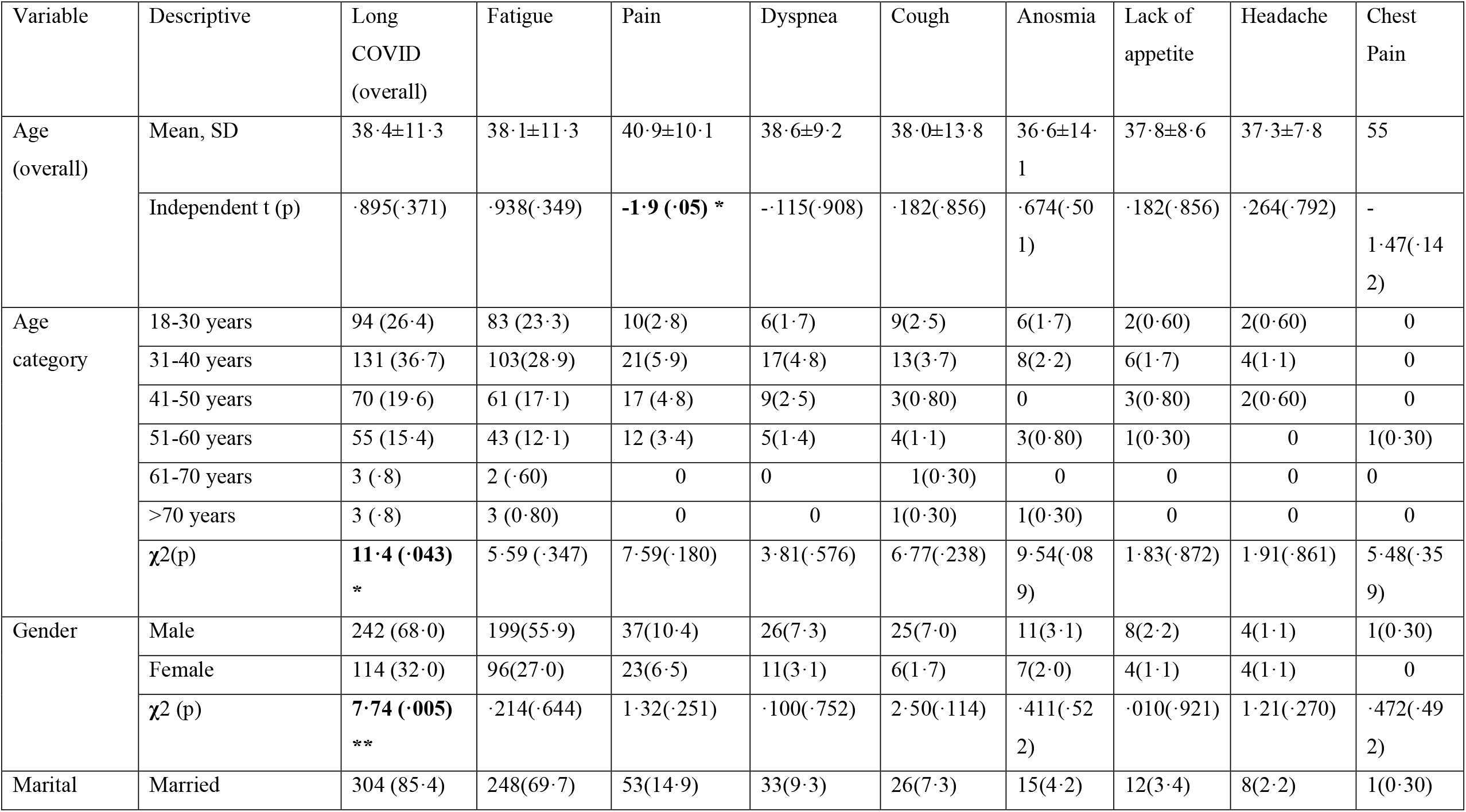
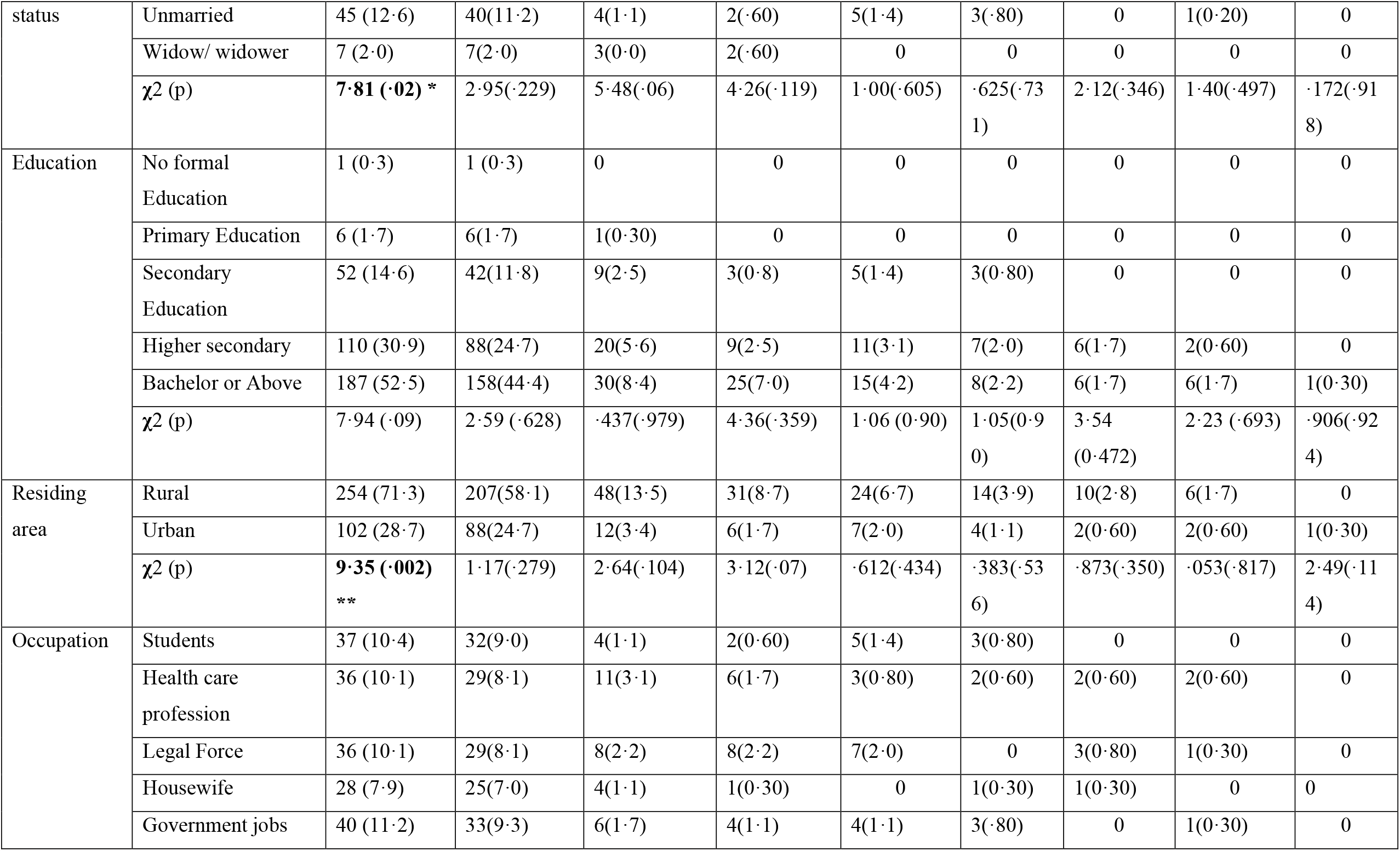
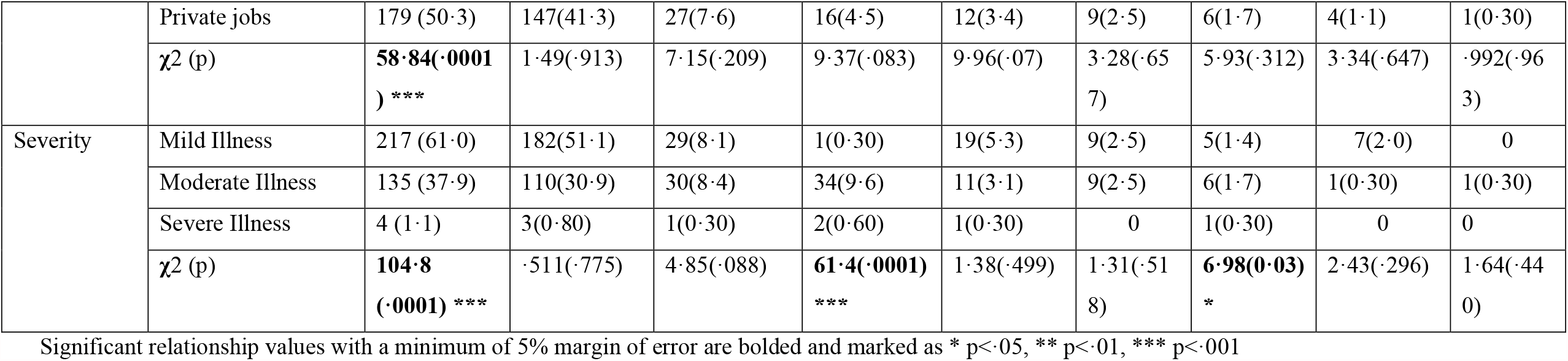
Relationship among socio-demographic variables and Long COVID symptoms of the COVID-19 survivors

**Table 3:**
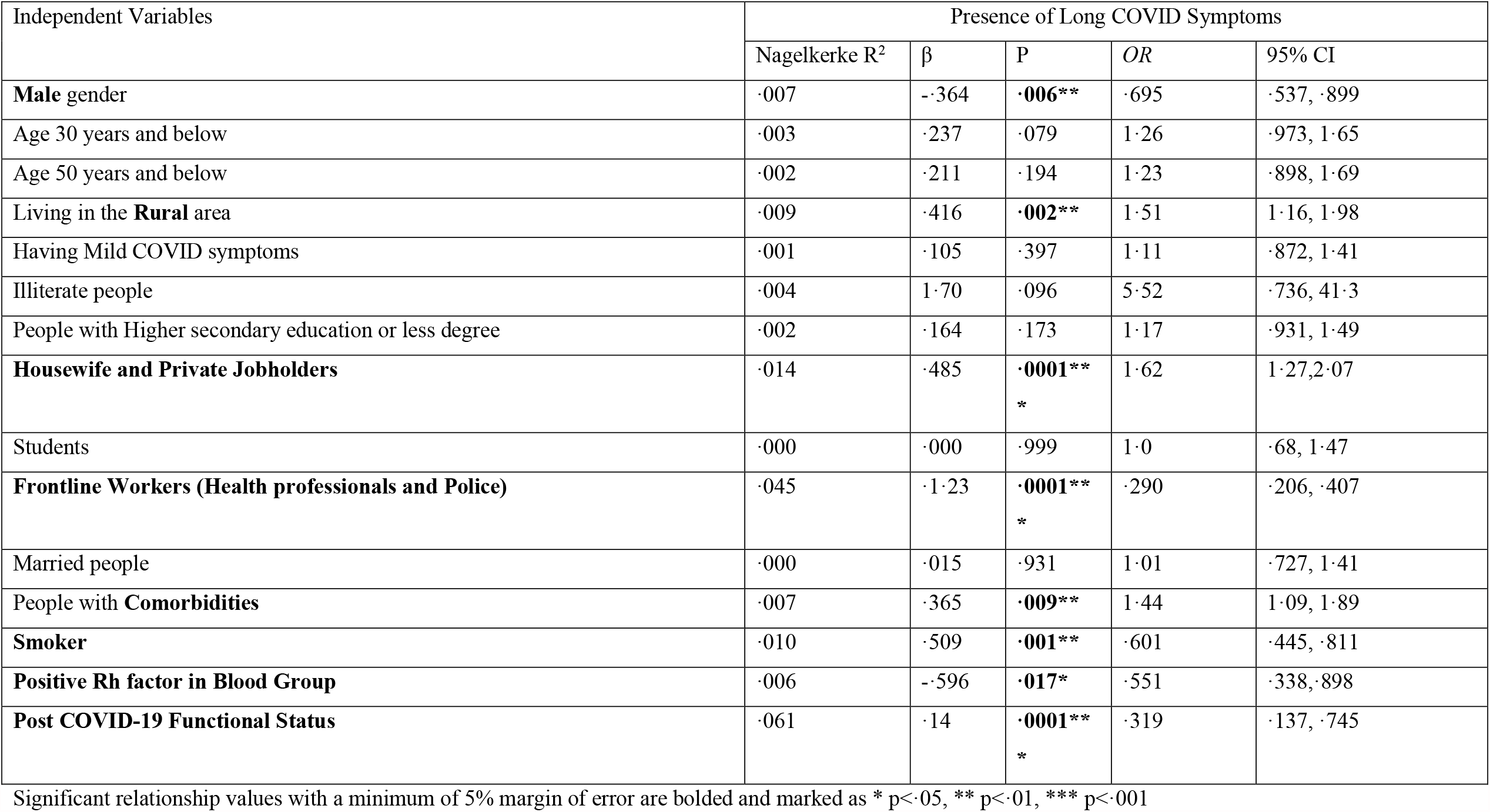
Factors associated with Long COVID symptoms

**Table 4:** Factors associated with limited function in people having Long COVID symptoms

| Independent Variables | Post COVID Functional Status Score |  |  |  |
| --- | --- | --- | --- | --- |
|  | R<sup>2</sup> | β | p | 95% CI |
| *Socio-demographics* |  |  |  |  |
| **Male** gender | ·017 | --.131 | **·014\*** | --.154, ·319 |
| **Female** gender | ·004 | ·064 | **·010\*** | ·194, ·262 |
| Age **50 years** and below | ·003 | -0·051 | **·0001\*\*\*** | ·845, ·900 |
| Rural people | ·000 | --.008 | ·752 | ·745, ·810 |
| Having **Mild COVID** symptoms | ·004 | --.065 | **·0001\*\*\*** | ·629, ·704 |
| *Long COVID Symptoms* |  |  |  |  |
| **Fatigue** | ·009 | ·094 | **·0001\*\*\*** | ·110, ·170 |
| **Pain** | ·031 | ·177 | **·0001\*\*\*** | --.018, ·011 |
| *Cardio-respiratory Functions* |  |  |  |  |
| **Resting Heart Rate** | ·099 | --.315 | **·0001\*\*\*** | -1·86, 1·85 |
| **Systolic Blood Pressure** | ·509 | ·714 | **·001\*\*** | -1·7, 2·7 |
| **Diastolic Blood Pressure** | ·569 | ·755 | **·001\*\*** | --.94, 3·1 |
| SpO2 | ·001 | ·012 | ·822 | --.92, 1·9 |
| **Inspiratory Lung Volume** | ·073 | ·271 | **·001\*\*** | -2·4, ·73 |
| **Expiratory Lung Volume** | ·485 | ·696 | **·001\*\*** | -1·26, 2·26 |
| **Vo2Max** | ·046 | ·213 | **·001\*\*** | -2·15, 2·53 |

Significant relationship values with a minimum of 5% margin of error are bolded and marked as \* p<·05, \*\* p<·01, \*\*\* p<·001

**Figure 3:**
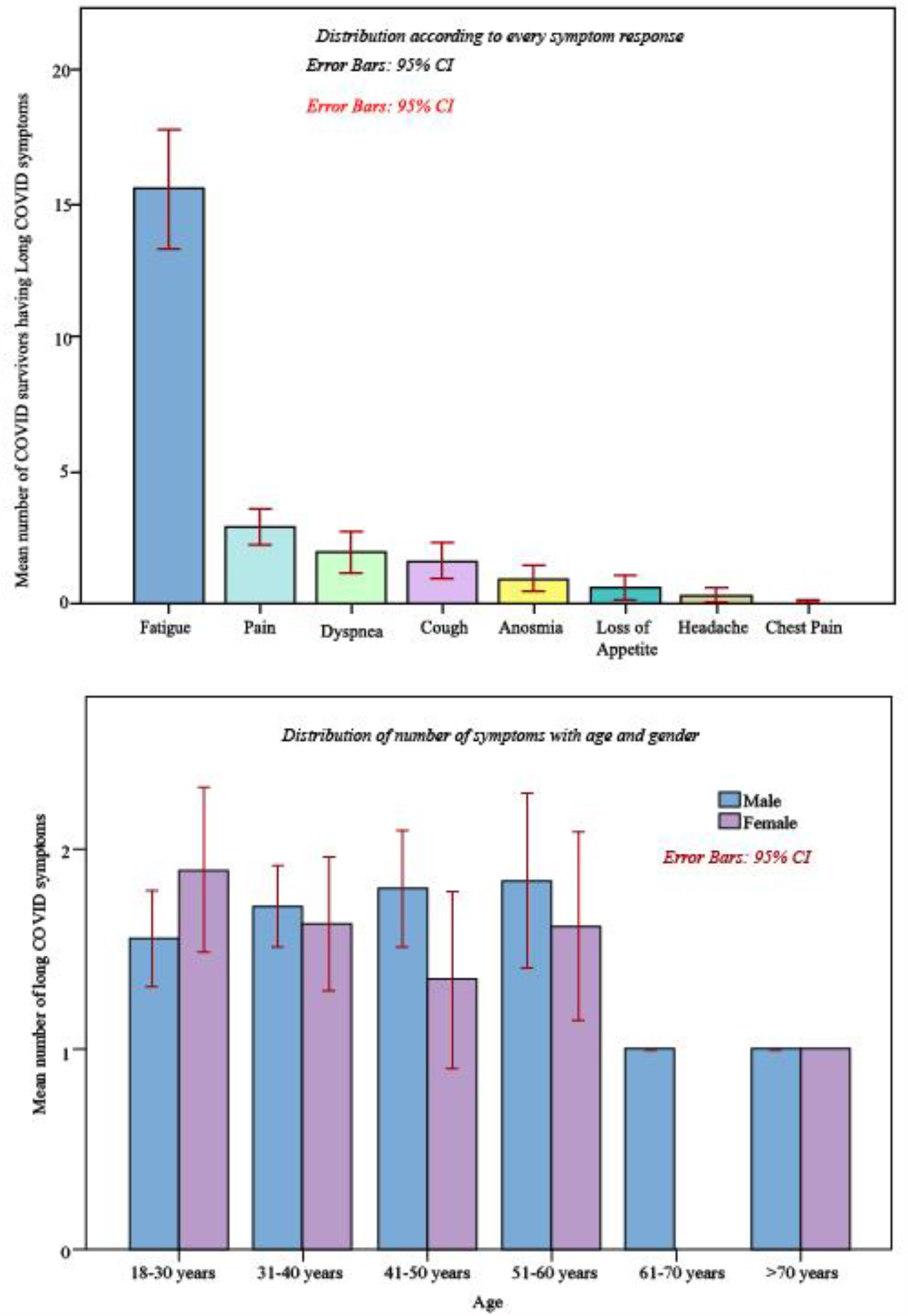
Distribution of Long COVID Symptom (LCS)

**Figure 4:**
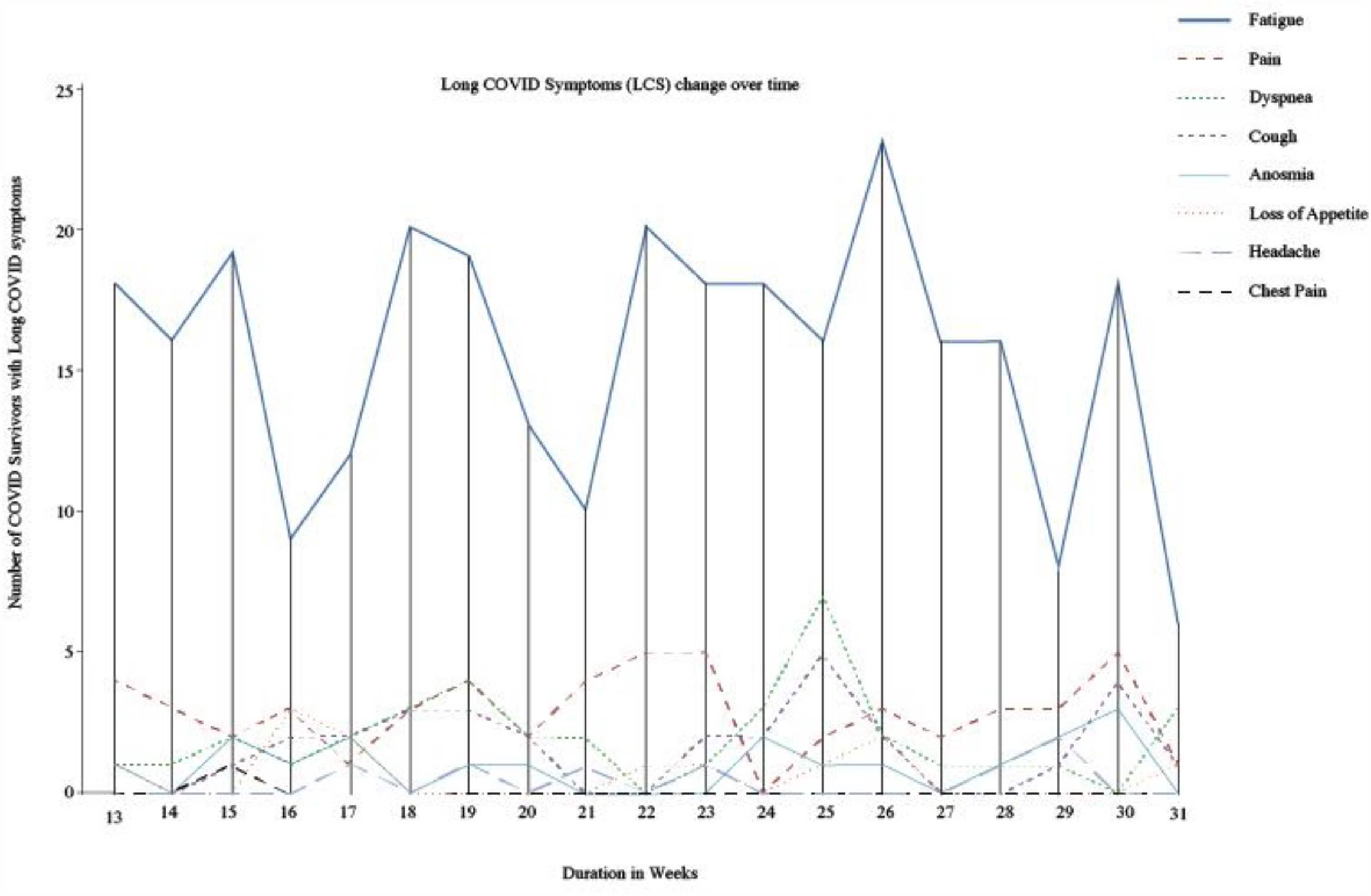
Relapsing Remittent pattern of Long COVID symptom (LCS)

**Figure 5:**
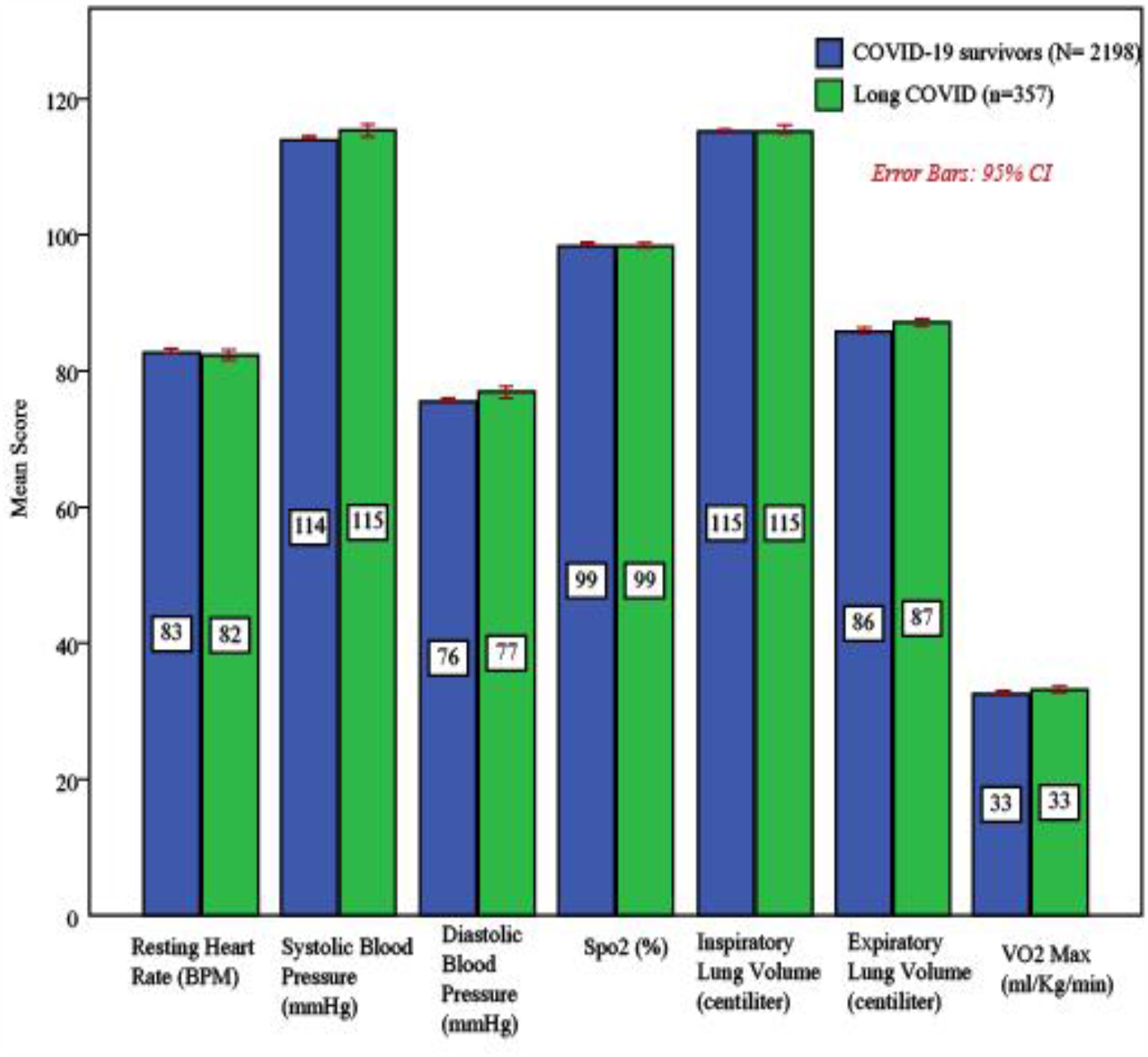
Comparison of Cardio-respiratory system variables between COVID-19 survivors and people with Long COVID symptoms (LCS)

**Figure 6:**
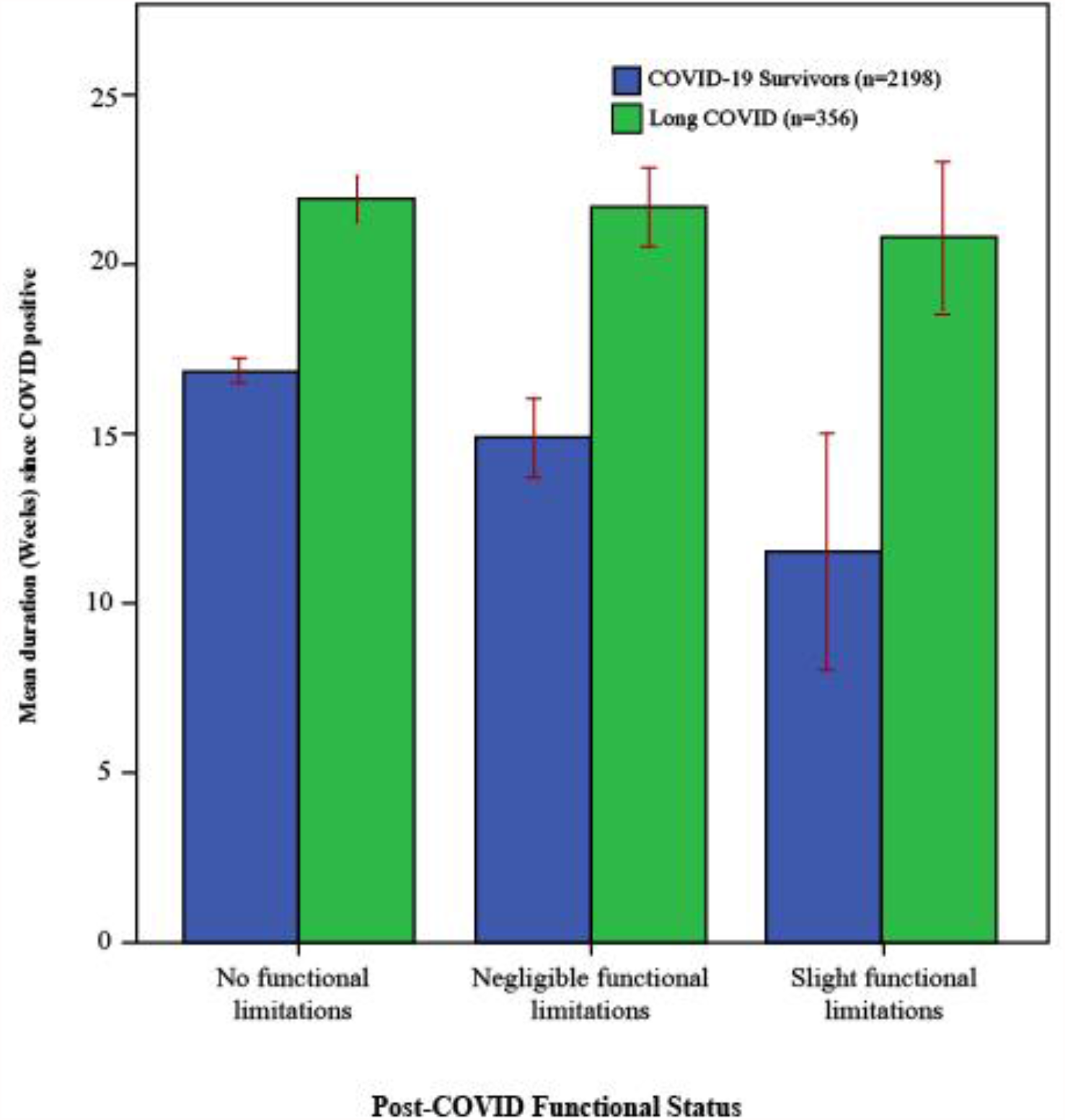
Comparison of Post-COVID Functional Status Scale between COVID-19 survivors and people with Long COVID symptoms (LCS) according to duration since COVID positive

**Figure 7:**
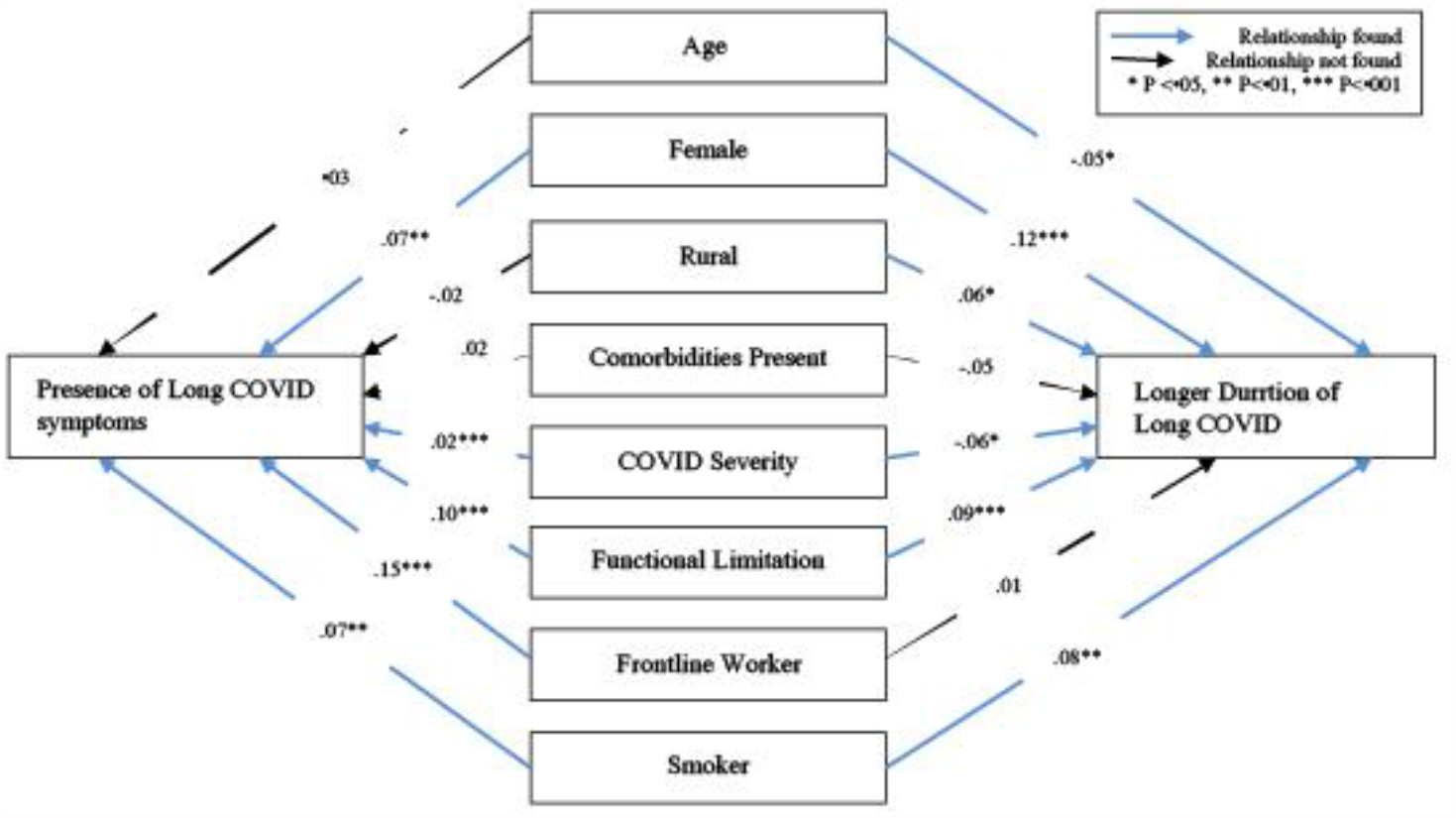
Structural equation model (SEM) for predictors of Long COVID Symptoms

### Patient and Public Involvement

Researchers invited the COVID-19 survivors from the list supplied by the government via telephone call before starting the face-to-face data collection. After preliminary screening, the eligible participants were listed separately and further contacted (Figure 1). Data was collected at the participant’s home or office with written consent. Participants had briefly demonstrated the study process, voluntary agreement and assured the confidentiality of their information by the data collectors during face-to-face interview. The patients declared their involvement and communication was willing, voluntary and had no objection to use the data for research purpose. The process of finding the contacts of the COVID-19 survivors, contacting them and perform data collection in this pandemic were permitted and approved by the appropriate authority of the Government of the People’s Republic of Bangladesh.

## Results

### Prevalence

From 14392 estimated samples, 13222 respondents were called by telephone.62.79% (8303) COVID survivors reported complete recovery and 27.23% (3920) stated that they faced difficulties when undertaking daily activities. The study had a reasonably good response rate of 56.8%. This study found that 22.5% (495) of COVID survivors had PACS at 4 weeks post diagnosis and 16.1% (356) had LCS at 12 weeks post diagnosis (Figure 1).

### Socio-demographic variables and health related information

The mean age of all ACS (n=2198) was 38.07 ± 11.4 years, survivors with PACS (n=495) 38.86±11.3 and those survivors with LCS (n=356) had mean age 38.42±11.3. The majority of the ACS were between 31 and 40 years of age 43.3% (718) and 30.4% (669) were aged 18-30 years. The male to female ratio of the respondents was 2.6:1. Respondents covered all eight administrative divisions in Bangladesh as follows: Dhaka 35.89% (789), Chittagong 8.50% (187), Rajshahi 8.37% (184), Sylhet 9.63% (212), Rangpur 9.13% (201), Barisal 10.62% (234), Khulna 8.04% (177) and Mymensingh 9.73% (214). There was a larger number of responses from rural areas 75.2% (1652). 85% (1868) of the participants were married and 50.1% (1101) were graduate or postgraduate. The most common occupation reported was that of private jobs, which comprised 55.1% (1212). Multiple response analyses found that the most common comorbidities for ACS were as follows: 22.7% had diabetes,25.6% had hypertension, 6% had heart disease, and 6.9% had pre-existing lung disease34.7% of the ACS had blood group B positive and other major distributions were O positive 23.4%, and A positive 23.1%. The detailed socio-demographic and health related information for participants is appended in Table 1.

### Symptom responses and duration

Figure 2 presents the results using the integrative post COVID symptoms model^7^and identifies that amongst the ACS13% (286) were asymptomatic. Amongst those with symptoms, the most common symptoms described were fever 76.6% (1683), fatigue 50.1% (1101), cough and upper respiratory tract symptoms 65.4% (1438), dyspnea 23.8% (523), pain 33.1% (727), ageusia 29.3% (644), headache 38% (836), and anosmia 43.9% (966). After four weeks, the major PACS were fatigue 84.8% (420), and pain 15.4% (76). After 12 weeks, LCS presented as fatigue 82.9% (295), cough and upper respiratory tract symptoms 8.7% (31), dyspnea 10.4% (37), pain 16.9% (60), chest pain .3% (1), ageusia3.4% (12), headache 2.2% (8) and anosmia5.1% (18). Fatigue was the prominent LCS. The number of LCS experienced by a COVID survivor ranged from 1 to 5 (Figure 3). The overall duration of LCS was 21.8±5.2 weeks. Duration of individual LCS varied in weeks as follows: fatigue 21.7±5.2; pain22.2±5.3; dyspnea22.7±4.9; cough and upper respiratory symptoms22.4±5.1; anosmia 22.6±6, ageusia20.6±5.5 and headache 22.5±5.7. There was a relapsing remitting pattern noted in the LCS from week 13 onwards to week 31, with the highest increments of LCS noted from 25th to 26th week (Figure 4).

### COVID related Information

According to WHO working group classification^24^, 63.2% (1390) of the ACS had mild COVID, 23.7% (520) had moderate COVID and .4% (8) had severe COVID-19. The ratio of the severity in ACS, PACS and LCS were mostly similar (Table 1).93.9% (2063) respondents opted to follow the advice of the government telemedicine team by resting at home, undertaking isolation and using advised medication. 5.7% (126) needed supplementary oxygen either at home or hospital, .2% (5) were admitted to hospital and required non-invasive ventilation and .2% (4) were admitted to a dedicated COVID Intensive Care Unit (ICU) with or without mechanical ventilation. Further detail of the COVID related information of ACS, PACS and LCS are appended in Table 1.

### Cardio-respiratory Function

The mean resting heart rate for participants with PACS and LCS was 82.5±6.9, and 82.4±6.7 beats per minute; systolic blood pressure was 115.8±8.9, 115.4±8.7 mm/Hg; diastolic blood pressure 77.3±7.6, 76.9±7.3 mm/Hg; SpO2 98.5±.6, 98.6±.7 %; inspiratory lung volume 1155±62, 1154±61 mL/min; expiratory lung volume 872±54, 871±56 ml/min; and Vo2Max 33.0±3.4, 33.1±3.3 respectively. However, figure 5 shows, no notable changes of mean in LCS compared to all COVID survivors.

### Post-COVID functional status (PCFS)

88.2 % (1939) of ACS had no residual functional limitation and74.9% (371) of PACS and 73.3% (261) of LCS also had no residual functional limitations. However, 20.2% (100) of PACS and 20.2% (72) of LCS reported negligible functional limitation and 4.8% (24) of PACS and 6.5% (23) of LCS reported slight functional limitations. Overall, the mean PCFS in PACS was 12.41±8.7 and in LCS was 12.14±8.8 in 0 to 100 scores. Compared to ACS, participants with LCS had higher mean scores in relation to progression of weeks (Figure 6), indicating that functional limitation is more progressive with time in LCS survivors compared to others.

### Relationship of LCS with Socio-demographics

Table 2 represents a segregated relationship of socio-demographic and health related factors with LCS in correlation model. Categorical age was associated with LCS (p<.05), whereas pain was associated separately with overall age of the population (p<.05). LCS were associated with gender (p<.01), marital status (p<.05), area of residence (p<.01), occupation (p<.001) and severity of COVID-19 (p<.001).

### Factors associated with LCS

Table 3 describes the binary logistic regression model to determine the factors associated with LCS. People living in rural areas had a relationship with LCS (*β*.41, p<.01. Other relationships included being a housewife and private employees (*β*.48, p<.001,), frontline worker (police and health professionals) (*β*.12, p<.001,), people with co-morbidities (*β*.365, p<.01), and smokers(*β*.50, p<.01).The PCFS was also found to be positively associated (*β*.14, p<.001).However, male gender had a reverse relationship with LCS (*β* -.36, p<.01), and the presence of a Rhesus positive factor in the blood group also had similar reverse relationship (*β* -.596, p<.05).

### Factors associated with limited function in LCS people

Table 4 shows the factors associated with limited functional scores in participants with LCS in a multiple linear regression model. The female gender was related with functional limitation (*β*.064, p<.05).Other linear associations were fatigue (*β*.094, p<.001), pain (*β*.17, p<.001), systolic blood pressure (*β*.71, p<.01), diastolic blood pressure (*β*.75, p<.01), inspiratory lung volume (*β*.27, p<.01), expiratory lung volume (*β*.69, p<.01) and Vo2Max (*β*.213, p<.01).A reverse relationship was found with males (p<.05), people aged below 50 years (p<.001) and people who had mild COVID-19 (p<.001).

### Prediction of LCS

Figure 7 predicts the presence of LCS and longer duration of LCS through the SEM. Goodness of fit indicates a satisfactory level of ‘good fit’ of assumptions for external validity (χ2 1302.88, df 29, p .001, χ2/df 44.92, Root mean square error of approximation (RMSEA).166 [90% CI .158, .174], Root mean square residual (RMR) 1.449, CFI .147, GFI .882). Presence of LCS was positively associated (*R*^*2*^.098) with frontline workers (police and health professionals) (*β*.15, p<.01), limited function (*β*.10, p<.001), being female (*β*.07, p<.001) smokers (*β*.07, p<.001), and the severity of COVID-19 (*β*.02, p<.001).A longer duration of LCS was positively associated (*R*^*2*^.041) with being female (*β*.12, p<.001), limited function (*β*.09, p<.001), smokers*(β*.08, p<.01), and people living in rural areas (*β*.06, p<.05). Interestingly, age has a reverse relationship with a longer duration of LCS (*β* -.05, p<.05).

## Discussion

In this study, the prevalence of LCS was 22.5% at 4 weeks and 16.1% at 12 weeks post diagnosis respectively. This is slightly higher than that reported by the UK study^11^ which reported the prevalence of PACS and LCS as 20% and 10% respectively. A recent study reported that the ongoing health issues of “fatigue or muscle weakness, sleep difficulties, and anxiety or depression.”^6^were experienced within a cohort of 1733 survivors discharged from hospital in China, when followed up at a six-month time point. The limitation of that study was that all participants had been treated as In-patients in hospital, unlike this research study, in which the majority of participants did not receive hospital treatment and remained at home during recovery. Our study adds valuable research knowledge to the gap in understanding the prevalence and nature of LCS in survivors, who have remained at home during their illness. Our study is larger than the two previous studies conducted in Bangladesh^13-14^ and provides new research knowledge on associated risk factors for LCS, in addition to identifying risk factors associated with a longer time length of these symptoms.

In this study, the most common symptom during the acute phase was fever, closely followed by fatigue and upper respiratory tract symptoms. This is consistent with literature that has reported similar symptoms.^25-26^The study found eight LCS, with fatigue being the most common symptom closely followed by muscle pain and dyspnea. In another study^27^, after three to nine months, 14% of individuals had fatigue problems. Most available literature unanimously reports fatigue^1,15,16^as the most common LCS. After this, many studies reportbreathlessness^28,1,6^ as the second most common LCS with other studies citing anosmia, cough and myalgia to also be common.^29^Augustin et al^30^, reported a study where non-hospitalized COVID-19 patients had more anosmia (12.4%) and ageusia (11.1%) than fatigue (9.7%) and shortness of breath (8.6%) over a four-to-seven-month recovery period.

This study reported that the majority of respondents had mild to moderate COVID-19 and approximately 93% decided to stay at home to recover without hospital treatment.62% (308) of respondents with LCS reported mild disease, which would not require hospitalization. There is a paucity of literature in this area, which perhaps reflects the challenge of how to collect and gather data from people who do not perceive that they are sick enough to require hospital treatment and also considering that Bangladesh is an under-resourced country and the hospital system is private so usually hospital admission and treatment costs money.

Figure 4 highlights the relapsing remitting nature of LCS reported over time between 13 to 31 weeks with maximum symptoms reported between 25 to 26 weeks, which is consistent with other research^1,5,7,12,17^. In figure 5, it is apparent that there was little difference noted between the cardio-respiratory function variables measured for PACS and LCS survivors. In terms of functional limitations, it is clear from Figure 6, that people with LCS had more functional limitations when compared with all ACS.

Overall, this study found that age (< 30 years); rural geographic residence; housewife role or private sector occupation; one or more comorbidities; smoking; a longer acute COVID recovery period, a positive Rhesus factor in the blood group and prior functional limitations were all predictive risk factors for LCS. To our awareness, this is the first study to identify specific risk factors for Long COVID within the general population and whilst the specific context is Bangladesh, we think that these risk factors warrant further investigation in all global community populations.

Previous studies on Long COVID in Bangladesh focused on urban areas and did not include rural regions, as this study did, which is now identified as a risk factor Whilst males (m) comprised the majority of respondents with LCS (69.0%), it was the females (f) who reported a greater proportion of LCS, (over all f = 25.2%, m=21.2%). This may be due to the overall higher incidence of COVID-19 diagnosed and reported by men in Bangladesh and the lack of sex disaggregated data in previous studies. The female sex was significantly associated with a longer duration of COVID symptoms. In addition, women reported slightly higher levels of fatigue, followed by pain, anosmia, and insomnia (f = 1.3%, m=0.3 %;). In contrast, men reported slightly higher levels of dyspnea, ageusia, cough and chest pain.

A significant positive association was found between gender and functional limitations. Women and those with cardio-respiratory comorbidity were more likely to develop functional limitations during the Long COVID period. In contrast, men and those with mild COVID symptoms were inversely associated with functional limitations.

The limitations of this study include that the sample was taken from twenty-four testing centres, and it is appreciated that people, who have attended these centres for diagnosis may not be truly representative of all communities within the Bangladeshi population. However, it will be possible for the team to contact and follow up on this cohort for longitudinal data in the future. In addition, the limited nature of resources meant that a proportion of potential participants could not be contacted due to system and educational constraints and the researchers had limited access to the electronic database. The LCS in this study were described by the survivors and screened by the data collectors, who were medical students. A clinical screening by healthcare professionals might reveal more in-depth symptom responses.

## Conclusion

In this study, the prevalence of LCS was identified to be 22.5% at 4 weeks and 16.1% at 12 weeks post diagnosis. In addition, the study identified some key predictors for the presence of LCS, in terms of associated risk factors and also risk factors associated with a longer duration of Long COVID illness. Further research is needed to gain more insight into these identified risk factors and what can be done to support communities affected by it.

## Data Availability

All data collected in this study is confidential to the study and will be shared anomalously or following ethical principle and made available to others according to the necessity of the study. Data will be available at a public repository

https://www.kaggle.com/kmamranhossain/long-covid-bd

## Data Availability

https://www.kaggle.com/kmamranhossain/long-covid-bd

## Data Availability

https://www.kaggle.com/kmamranhossain/long-covid-bd

## Contributors

MAH, KMAH, KS, ZU, LMW, VR, MS, RS, MSH, MFK, RF, SR, MSA, SKC, MAH, IKJ conceptualised the study. MAH, KMAH, LMW, VR, RS, SR, MSA, IKJ curated and collated the data. MAH, KMAH, KS, ZU, LMW, VR, MS, RS, MSH, MFK, RF, SR, MSA, SKC, MAH, IKJ did validation. KMAH, KS, ZU, LMW, VR, MS RS, SR, MSA visualised the data. MAH, KMAH, KS, ZU, LMW, VR, MS, RS, MSH, MFK, RF, SR, MAH, IKJ contributed to the model development. MAH, KMAH, KS, MS, RS, RF, SR, MSA, IKJ did the formal analysis. MAH, KMAH, KS, LMW, VR, RS wrote the manuscript. KS, ZU, LMW, VR, MS, MSH, MFK, RF, SR, MSA, SKC, MAH, IJK reviewed the manuscript. All author had full access to all the data in the study and the corresponding author had final responsibility for the decision to submit for publication.

## Ethical approval

Ethical permission was obtained from the Institutional Review Board at the Institute of Physiotherapy, Rehabilitation, and Research (Ethical review committee at Bangladesh Physiotherapy Association) on September 17, 2020 (BPA-IPRR/IRB/17/09/2020/028) and the study was registered at World Health Organization (WHO)Primary Clinical trial registry platform (CTRI/2020/09/028165) on 30/09/2020 with the title “Symptoms presentation among the COVID-19 survivors in Bangladesh”. Written approval for data collection obtained from the Directorate General of Health Services (DGHS) of the Ministry of Health and Family Welfare in Bangladesh. Verbal consent was obtained during the initial telephone call and written consent was obtained at interview. The principles of the Helsinki Declaration^19^ were followed throughout the research to ensure confidentiality, ethics and privacy.

## Data Sharing

All data collected in this study is confidential to the study and will be shared anonymously or following ethical principle and made available to others according to the necessity of the study. Data will be available at a public repository. (https://www.kaggle.com/kmamranhossain/long-covid-bd).

## Acknowledgments

We thank all individuals who participated in this study and their families.

